# Feasibility of high-density surface electromyography for the detection of neuromuscular disorders in children

**DOI:** 10.1101/2025.06.23.25329951

**Authors:** Eduardo Martinez-Valdes, Ignacio Contreras-Hernandez, Ragul Selvamoorthy, Francesco Negro, Andrew Lawley

**Affiliations:** School of Sport, Exercise and Rehabilitation Sciences, College of Life and Environmental Sciences, University of Birmingham, Birmingham, UK; Department of Clinical and Experimental Sciences, Universita degli Studi di Brescia, Brescia, Italy; Department of Neurophysiology, Birmingham Children’s Hospital, Birmingham, UK

**Author notes:** **Corresponding authors:** Dr Eduardo Martinez-Valdes, Assistant Professor in Physiotherapy, School of Sport, Exercise and Rehabilitation Sciences, College of Life and Environmental Sciences, University of Birmingham, Edgbaston B15 2TT, United Kingdom,; Dr Andrew Lawley, Consultant Clinical Neurophysiologist, West Midlands Training Programme Director for Clinical Neurophysiology, Birmingham Women’s and Children’s NHS Foundation Trust. Authors contributed equally to this work (Ignacio Contreras-Hernandez, Ragul Selvamoorthy).

**Keywords:** Motor unit, motor neuron, pediatric, myopathy, neuropathy

## Abstract

**Introduction/Aims:** Diagnosing neuromuscular disorders in children is challenging. Concentric needle electromyography (CNEMG) is the standard for electrophysiological assessments but has limitations in paediatric populations. High-density surface electromyography (HDsEMG) offers a non-invasive alternative with better spatial resolution, allowing the identification of full motor unit (MU) firing dynamics. This study assessed the feasibility of HDsEMG MU decomposition in children and explored parameters that differentiate neuropathy, myopathy, and normal findings.

**Methods:** One hundred children (9.1±5.1 years) underwent CNEMG followed by HDsEMG. EMG signals were decomposed into individual MU spike trains, and MU yield, as well as firing properties (mean discharge rate (MDR), discharge rate variability (DRV), were analysed across diagnostic groups. Furthermore, MU action potential parameters obtained from CNEMG (MU amplitude and duration) were correlated against those obtained from HDsEMG.

**Results:** MUs were reliably identified in 86.0% of children, with an average of 7 (4.2) MUs per participant. Among MU firing parameters, DRV was significantly higher in children with myopathy (p=0.005). Additionally, MU duration from HDsEMG correlated significantly with CNEMG values (r=0.31) and successfully discriminated myopathy from normal and neuropathic groups (p=0.02).

**Discussion:** HDsEMG MU decomposition is feasible in children with neuromuscular disorders, providing valuable insights into MU firing and MU action potential properties. This technique has the potential to improve diagnosis and monitoring of paediatric neuromuscular conditions.

## INTRODUCTION

Neuromuscular disorders may present with a range of symptoms in infants and young children. Electrodiagnostic studies can be a useful diagnostic tool to help localise a disorder affecting the muscle, neuromuscular junction, peripheral nerve or anterior horn cell ^1,2^.

Identification of myopathy can be challenging in young children, with a wide range in reported diagnostic sensitivity between 10% and 86% ^3-6^. This may reflect variations in study populations, electromyography (EMG) analysis techniques and operator experience. Quantitative motor unit action potential (MUAP) analysis may improve diagnostic sensitivity ^3^, although limitations still exist. Firstly, needle EMG examination may be uncomfortable for young children ^7^. Secondly and related to the first point, EMG examination in young children typically involves fewer needle insertions than in adults, which may miss patchy distribution of muscle involvement. Thirdly, normal MUAPs in infants have shorter durations compared to adults, reflecting smaller diameter of muscle fibres and endplate zones ^8^. This can make it challenging to distinguish between pathological and normal MUAPs in infancy, even with quantitative MUAP analysis techniques.

Techniques to improve electrodiagnostic studies in young children would ideally be non-invasive, allow increased sampling both in terms of the number of muscles that can be studied but also the area of individual muscles, and potentially allow analysis of parameters other than those related MUAP morphology. High-density surface electromyography (HDsEMG) is widely used in physiological sciences, sports and neuroscience research. Previous investigations from a single centre reported inconclusive results with this technique in children with neuromuscular disorders ^9-11^. Despite the significance of this pioneering research and its considerable potential in pediatric populations, it was unfortunately not pursued further in subsequent investigations. However, there have been considerable recent advances in HDsEMG, particularly motor unit identification with automatic EMG signal decomposition algorithms ^12-14^ and the reliability and reproducibility of the technique has been demonstrated in adults ^15^.

The aims of this study are, firstly, to explore the feasibility of recording HDsEMG in children in a clinical setting, and secondly, to explore the potential diagnostic utility of HDsEMG by comparing motor units decomposed from HDsEMG recordings to those recorded with needle EMG.

## METHODS

### 2.1 Participants and procedures

This was a prospective study performed over a 12-month period between May 2023 and May 2024 in the Clinical Neurophysiology department at Birmingham Children’s Hospital. All children (age 0 to 16 years) referred for electrodiagnostic studies with a clinical question of an underlying neuromuscular disorder were eligible for inclusion. Exclusion criteria included children with skin lesions or a history of allergy to materials used for recording HDsEMG. Informed written consent was taken to perform HDsEMG recordings in addition to standard electrodiagnostic studies. To explore patient experience, all children who could understand necessary instructions were asked to complete a revised Faces Pain Scale ^16^ for both needle EMG and HDsEMG. Ethical approvals were granted from University of Birmingham research governance department and from the NHS Health Research Authority (IRAS ID: 319384).

### 2.2. Electrophysiology – Concentric needle EMG

Nerve conduction studies and concentric needle EMG (CNEMG) examination was performed by the same investigator (AL). No sedation was used. Temperature was checked and warmed to at least 30°C in lower limbs and 32°C in upper limbs. Muscle selection for CNEMG examination was guided by presenting symptoms, patient age and tolerance of the test, with tibialis anterior and biceps brachii most frequently examined. EMG was performed using a 30G Ambu Neuroline Concentric needle electrode and recorded with Sierra Summit software version 4 (Cadwell Inc, Kennewick, WA, US), with filter settings of 5Hz-10kHz. Multi-MUAP analysis was performed offline using automated software. At least 5 individual MUAPs were required to confirm a MUAP was unique and MUAPs with amplitude less than 100µV were excluded from analysis. MUAP analysis was performed at 200µV/Div and automated placement of cursors was accepted unless clearly incorrect. Overall diagnostic conclusion was based on combination of qualitative and quantitative assessment, particularly measurement of MUAP duration with multi-MUAP analysis. CNEMG findings were classified as normal, myopathic or neuropathic.

### 2.3. Electrophysiology - HDsEMG

HDsEMG was recorded using a 64-electrode grid, with 4 or 8mm interelectrode distance (IED) depending on muscle size, placed over the muscle of interest. HDsEMG was sampled at 2048Hz and converted to digital data by a 16-bit analogue to digital converter (64-channel EMG amplifier Sessantaquattro, OT Bioelettronica, Torino, Italy). Electrodes were attached using conductive paste to create good contact between skin and electrodes. Recordings were made using OT Biolab+ software and later analysed with Matlab (The Mathworks Inc., Natick, Massachusetts, USA). HDsEMG was initially recorded during spontaneous movements, in the same way that needle EMG recordings were obtained. Subsequently, where children were old enough to follow simple instructions, a videogame EMG biofeedback app (OT Bioelettronica Planes) was used to standardise level of muscle contraction. The app uses the amplitude of the envelope of the interference pattern during maximal volitional contraction of the muscle. Therefore, the child was first asked to perform three maximal voluntary contractions, and the peak EMG value (50ms window) was used as a reference for submaximal contractions. Following the maximal contraction assessment, the child was encouraged to maintain a 20s contraction at 20% of the maximal EMG. This procedure allowed stable HDsEMG signals, without the need for additional dynamometers or force transducers.

### 2.4 HDsEMG Signal analysis

The HD-sEMG signals recorded during the isometric contractions were decomposed into motor unit spike trains with an algorithm based on blind source separation, which provides automatic identification of multiple single motor units ^14^. Single motor units were assessed for decomposition accuracy with a validated metric (Silhouette, SIL), which was set to > 0.80. SIL is a normalized measure of the relative height of the peaks of the decomposed spike trains with respect to the baseline noise and it related to the sensitivity of the decomposition ^17^. In this study, the SIL value was set at a lower threshold compared to previous studies in healthy adults since data in pathological individuals may exhibit unphysiological firing patterns that could be potentially discarded by the decomposition algorithm if a high SIL threshold is used^17^.

Unlike CNEMG, which analyzes MUAP morphology based on the average MUAP from a minimum of five firings, HDsEMG allows for the decomposition of signals throughout the entire contraction, enabling assessment of motor unit firing patterns for the full duration of unit activity. Discharge times of the identified motor units were converted into binary spike trains, and firing properties were analysed using instantaneous firing rate profiles (calculated as the inverse of the interspike interval: 1/interspike interval). The mean discharge rate and discharge rate variability (quantified as the coefficient of variation for discharge rate [CVDR], defined as SD discharge rate /mean discharge rate × 100) were assessed. These values were computed at the time point when participants were able to maintain a stable EMG level during the contraction. In addition, parameters such as recruitment and de-recruitment threshold were calculated. Recruitment and de-recruitment thresholds were defined as the %EMG level where motor units began and ceased firing action potentials during the contraction respectively.

Since HDsEMG decomposition algorithms can introduce errors in firing pattern identification—such as missing pulses leading to abnormally long interspike intervals—all motor unit discharges were visually inspected and corrected when necessary. Additionally, merged motor units (where the algorithm erroneously classified two distinct units as one, combining their firing activity) were identified and excluded from analysis when detected ^17^. However, the quality control process was less conservative than in studies on healthy individuals, as pathological firing patterns may naturally include irregular spiking intervals or prolonged pauses in action potential firing ^18^.

Besides the analysis of motor unit firing data, HDsEMG MUAP morphology parameters such as MUAP amplitude and duration were assessed and compared between HDsEMG and CNEMG, given that these values are commonly used in clinical practice. HDsEMG MUAP amplitude values were assessed by calculating the average root mean square and peak to peak values of all EMG channels of the electrode grid in bipolar-single differential derivation (59 channels). The duration of HDsEMG MUAPs was estimated by calculating the time lag between the beginning of the MUAP rise (depolarisation) and its peak. This value was also averaged across the 59 bipolar channels of the electrode grid.

All motor unit data was recorded, analysed, and reported according to the consensus for experimental design in electromyography: single motor unit matrix ^17^.

### 2.4 Statistical analysis

Demographic variables (e.g., age, weight), CNEMG motor unit characteristics (number of identified units, mean duration [ms], mean amplitude [µV], and polyphasic percentage [%]), and HDsEMG motor unit parameters (mean discharge rate, CoV_ISI, MUAP amplitude, and duration) were compared across groups (normal, myopathy, and neuropathy) using one-way analysis of variance (ANOVA). If ANOVA results were significant, pairwise post-hoc analysis were made with a Tukey test.

Differences in discomfort levels between CNEMG and HDsEMG (assessed using the Happy Faces scale) were analyzed using a paired t-test. Associations between CNEMG and HDsEMG MUAP parameters (amplitude and duration) were examined using the Pearson correlation coefficient (r) and linear regression.

Motor unit firing parameters were compared across groups using data from the entire study population (N = 96, see participants section below). However, comparisons of MUAP amplitude and duration were restricted to children who used an 8mm inter-electrode distance (i.e.d) grid (N = 78), as variations in inter-electrode spacing can influence these parameters.

## RESULTS

### 3.1 Participants

110 potential participants were identified, with 100 consenting for HDsEMG recordings. Participant age ranged from 11 days corrected age to 16 years, with tibialis anterior most examined. Diagnostic findings of the neurophysiology study were normal for 62 participants, with 19 having myopathic EMG abnormalities, 15 having neuropathic EMG abnormalities and 4 showing mixed or inconclusive findings. These 4 participants were not included in the analysis. Full participant demographics and CNEMG results for 96 participants are presented in table 1.

**Table 1.** Participant characteristics and CNEMG results.

|  | NORMAL | MYOPATHY | NEUROPATHY | P-VALUE |
| --- | --- | --- | --- | --- |
| Number of participants | 62 | 19 | 15 | ----- |
| Sex | 40 M/22 F | 13 M/6 F | 7 M/8 F | ----- |
| Age (y) | 9.6 (5.2) | 7.3 (4.2) | 9.0 (5.4) | 0.23 |
| Height (cm) | 138.2 (36.4) | 124.3 (25.5) | 132.4 (29.6) | 0.32 |
| Weight (kg) | 41.9 (25.8) | 30.5 (15.8) | 35.7 (21.1) | 0.18 |
| Temperature (C) | 31.4 (1.3) | 31.8 (1.4) | 31.8 (1.1) | 0.32 |
| Examined muscle | BB=11, TA=51 | BB=3, TA=16 | BB=1, TA=11, AP=1, ED=2 | ----- |
| **Motor Units (N)** | **14.0 (6.3)** | **11.3 (5.7)** | **7.9 (4.4)** | **0.02** |
| **Mean duration (ms)** | **8.6 (1.0)** | **6.6 (1.5)** | **13.4 (4.1)** | **<0.001** |
| **Mean amplitude (uV)** | **512.8 (159.0)** | **409.9 (186.5)** | **1179.6 (750.7)** | **<0.001** |
| **Polyphasic (%)** | **4.2 (5.3)** | **19.2 (22.4)** | **16.7 (22.6)** | **<0.001** |

BB = Biceps Bracchi, TA = Tibialis anterior, AP = Adductor pollicis brevis, ED = Extensor digitorum

### 3.2 Participant reported discomfort

Revised faces pain scales were completed by 80 participants (age 11.0 (3.8) years), with a score of 0 indicating no pain and 10 indicating maximum pain. Pain scores were significantly lower for HDsEMG compared with CNEMG examination (0.45 (0.95) *vs* 4.03 (2.5), *p<0.001)*. Only 1 participant reported a preference for CNEMG examination but reported low level of discomfort for both procedures (4 *vs* 2). 64 participants reported no discomfort with HDsEMG. Where discomfort was experienced, this was related to either electrode removal or continuous muscle contraction. Further details are provided in **figure 1**.

**Figure 1.**
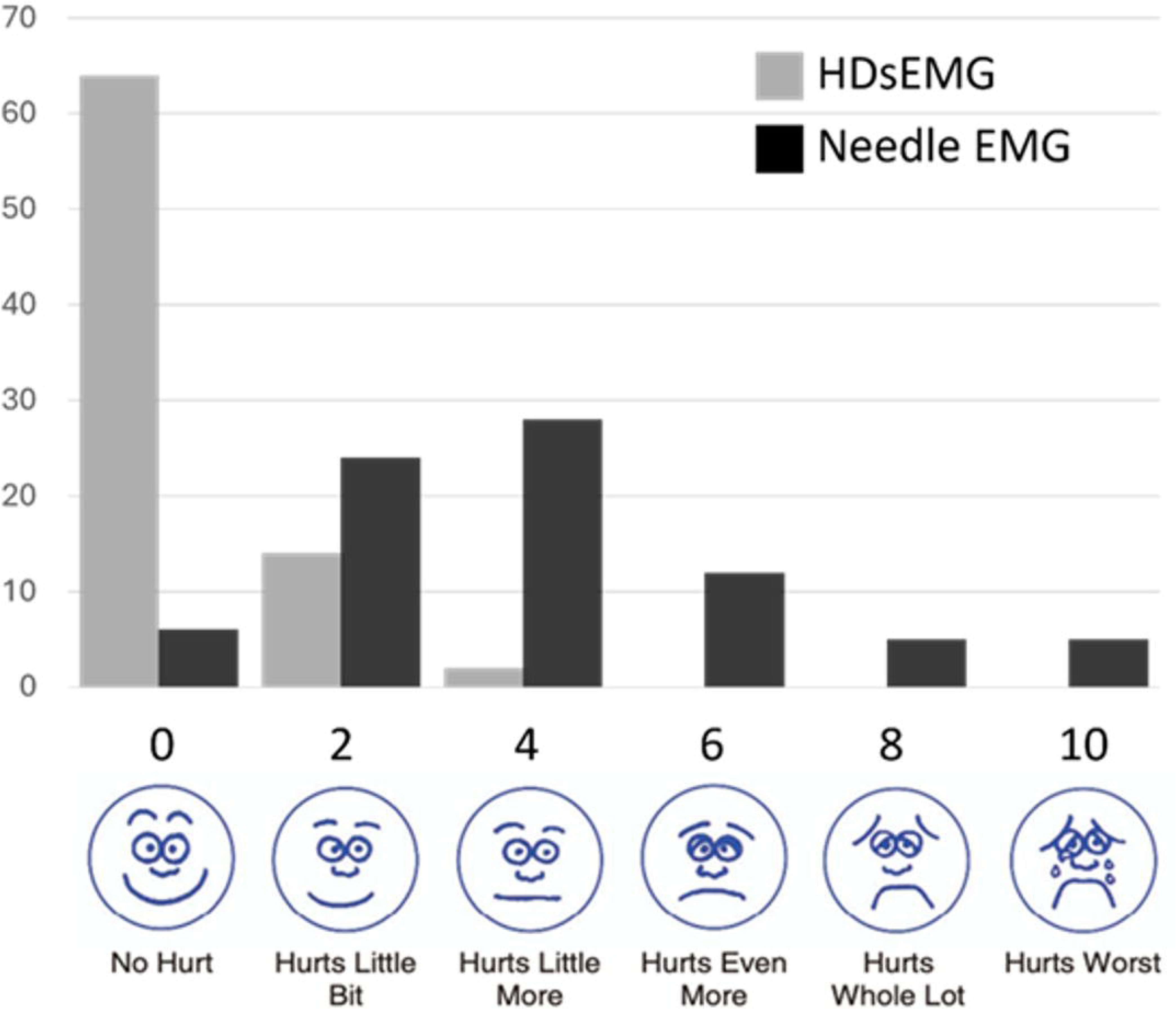
Revised faces pain scale response distribution for HDsEMG and CNEMG (n=80).

### 3.3 HDsEMG decomposition – motor unit identification

A total of 101 HDsEMG recordings were made in 100 participants. 18 recordings were made with 4mm IED; the remainder with 8mm IED. Following HDsEMG decomposition, motor units with a full firing pattern (i.e., motor units active for more than 25% of contraction time) could be identified on 86 children. From these children, 7 (4.2) motor units could be identified on average per-participant. The ability of the decomposition algorithm to detect motor units varied between normal, myopathic and neuropathic groups, with the myopathy group being the group with the lower number of motor units identified (4.2 (5.4)) versus normal (7.4 (4.7)) and neuropathic (7.0 (3.2)), who had similar number of identified units (condition effect p=0.034). Furthermore, the ability of the algorithm to detect motor units was associated with the age of the participants (r=0.447, p<0.001) as the number of observed units increased with age (**Figure 2**).

**Figure 2.**
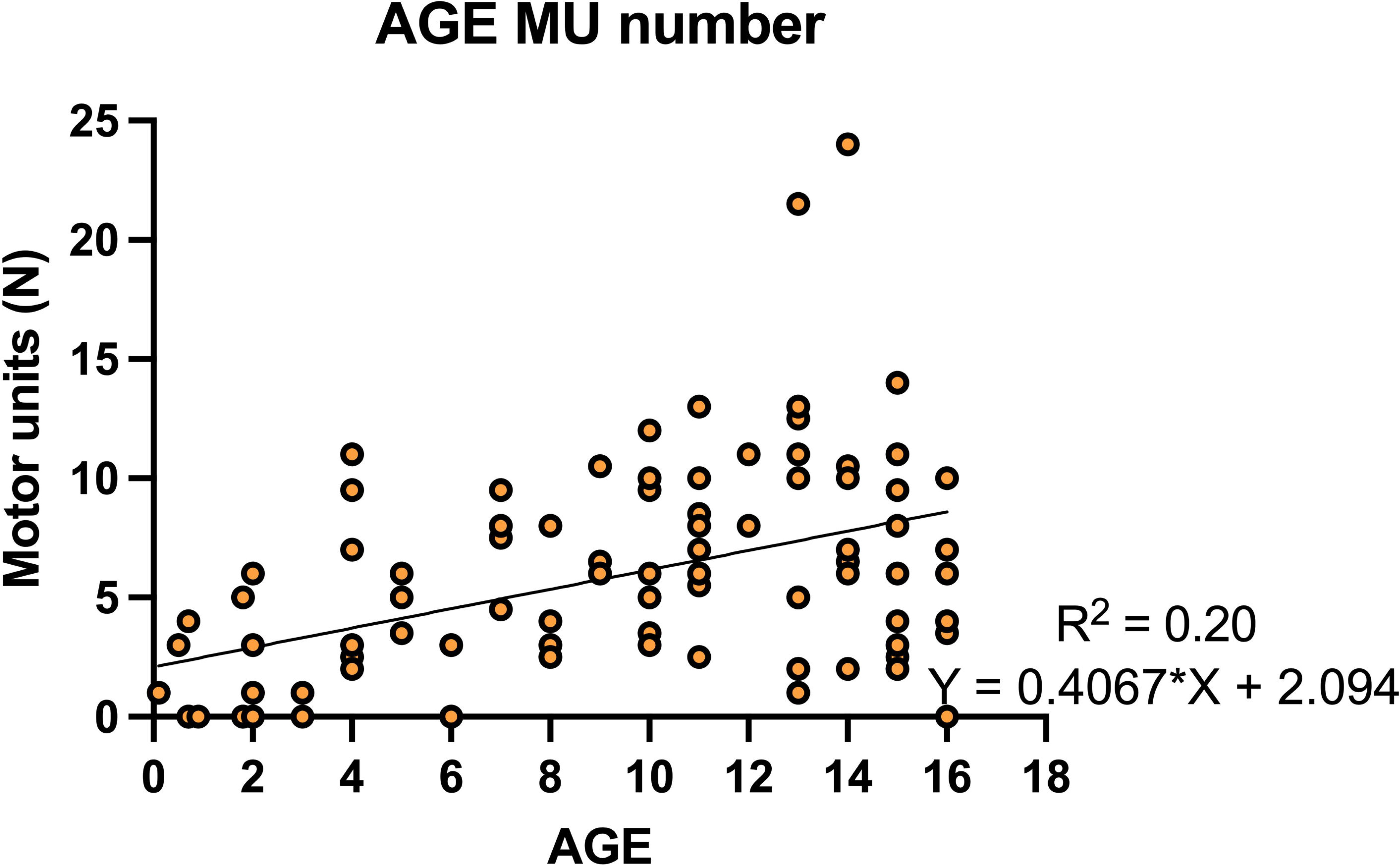
Association between participant age and number of motor units identified.

### 3.4 HDsEMG motor unit firing analysis

Motor unit recruitment and de-recruitment thresholds were comparable across groups (F = 0.728, p = 0.486 and F = 0.750, p = 0.476, respectively), indicating that similar populations of motor units were assessed. Specifically, recruitment thresholds were 22.5% (16.7) MVC EMG for the normal group, 24.2% (15.5) MVC EMG for the myopathic group, and 21.6% (12.3) EMG for the neuropathic group. Mean discharge rate showed no significant differences between groups (F = 0.402, p = 0.670). However, the coefficient of variation for discharge rate (CVDR) was significantly higher in the myopathic group compared to both the normal and neuropathic groups (F = 5.7, p = 0.005). Motor unit firing results are presented in **Figure 3**.

**Figure 3.**
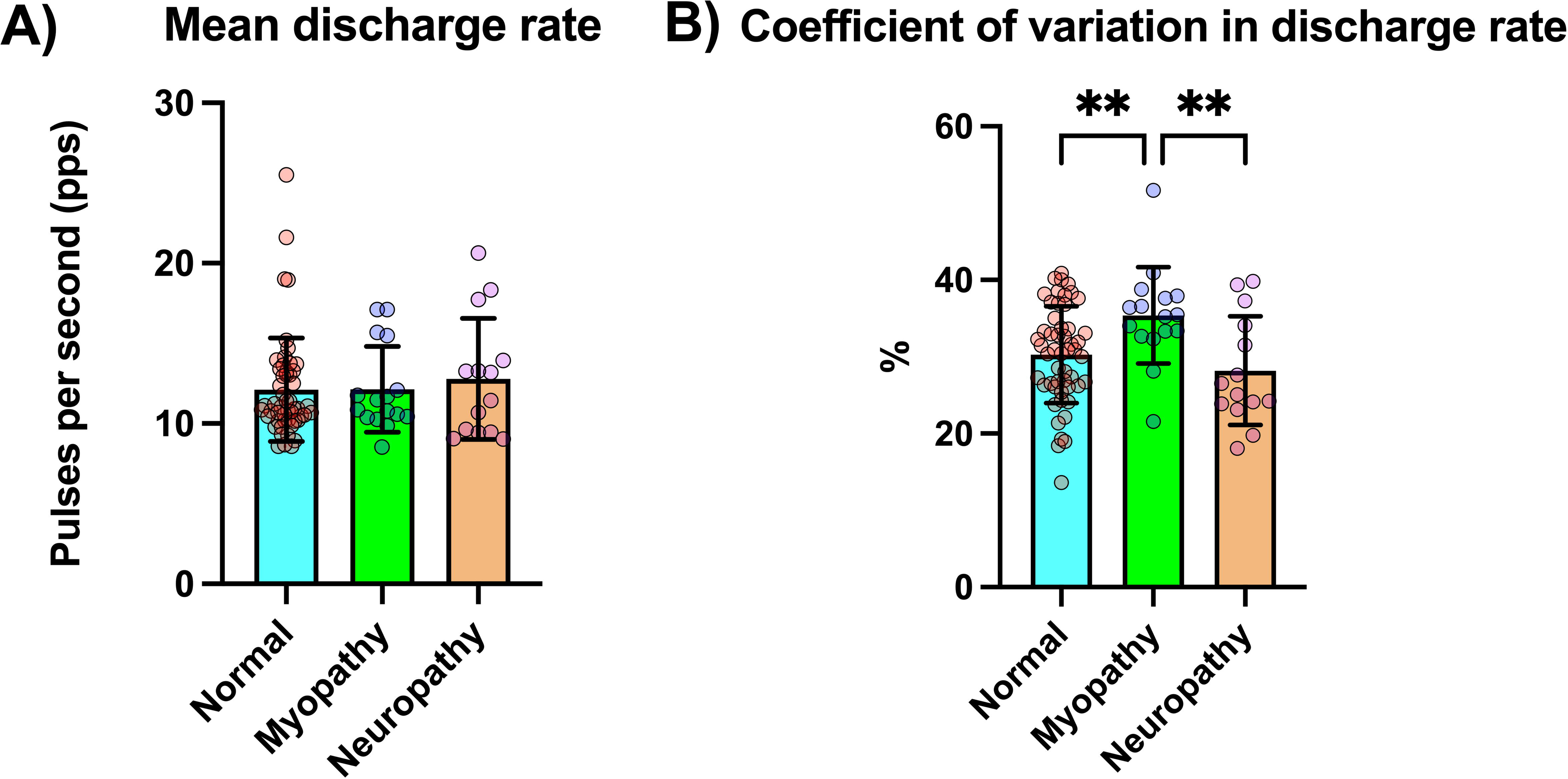
Motor unit firing results across the normal, myopathy and neuropathy groups. A) Mean motor unit discharge rate, B) discharge rate variability quantified as the coefficient of variation in discharge rate. P<0.001**

### 3.5 Association between HDsEMG and CNEMG parameters

MUAP amplitude values obtained from CNEMG recordings showed no significant correlation with HDsEMG MUAP peak-to-peak values (r = 0.20, p = 0.096). However, MUAP duration from CNEMG analysis was significantly correlated with MUAP half-duration values from HDsEMG (r = 0.31, p = 0.0086). **Figure 4** illustrates the association results between CNEMG and HDsEMG MUAP amplitude and duration. While HDsEMG MUAP amplitude values did not distinguish between groups (F = 1.038, p = 0.36), HDsEMG MUAP duration values were significantly shorter in the myopathic group compared to the normal and neuropathic groups (F = 4.024, p = 0.02). Comparisons of HDsEMG parameters across groups are presented in **Figure 5**.

**Figure 4.**
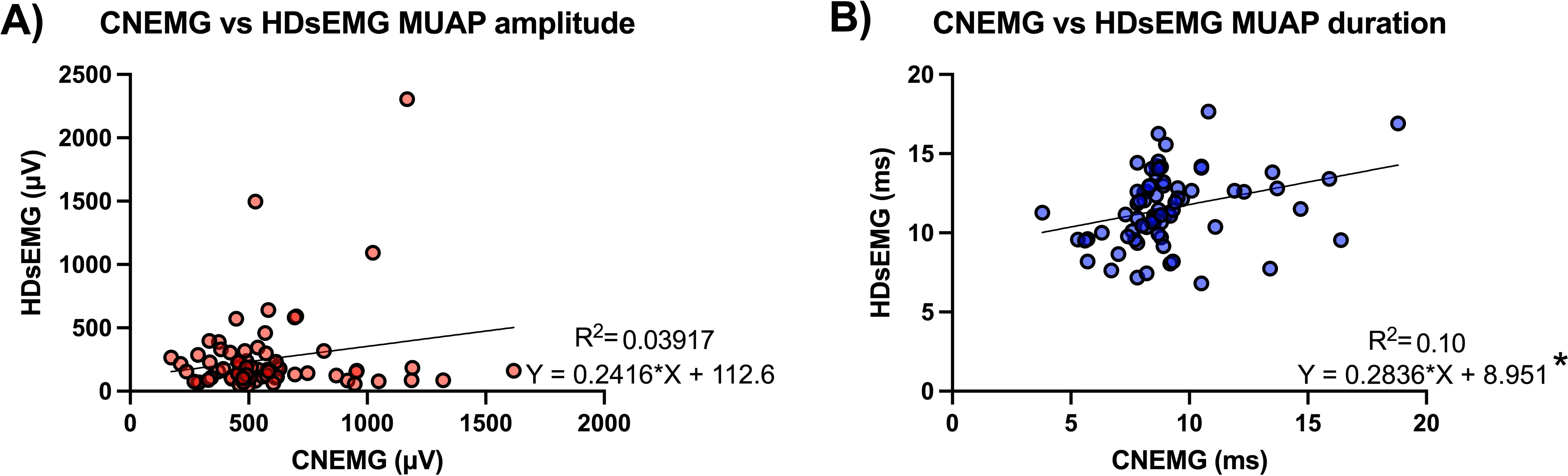
Association between MUAP parameters obtained from CNEMG and HDsEMG. A) HDsEMG versus CNEMG MUAP amplitude. B) HDsEMG versus CNEMG MUAP duration. Significant association (p=0.0086)*

**Figure 5.**
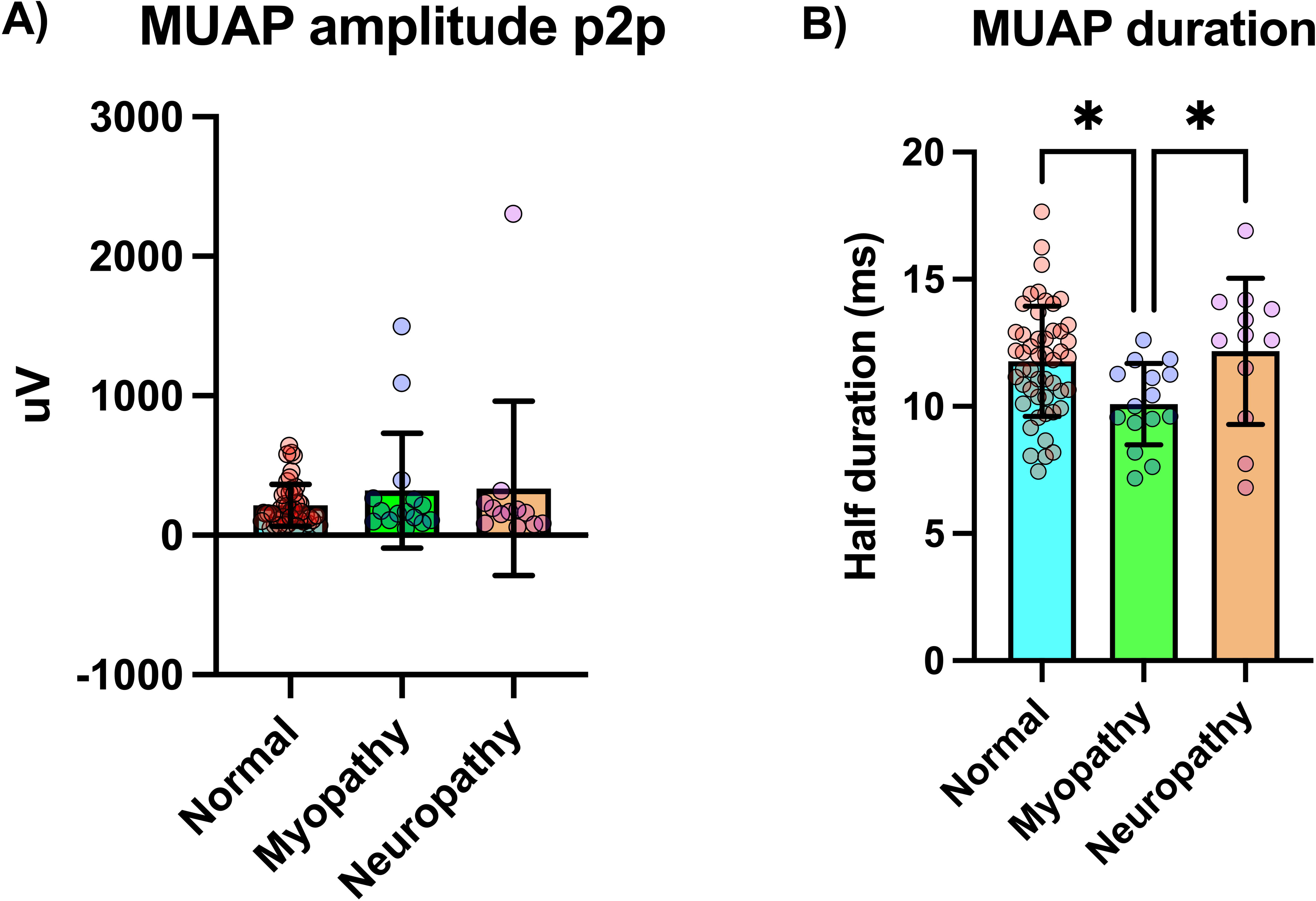
MUAP parameters from HDsEMG recordings compared across groups. A) MUAP amplitude values (peak-to-peak MUAP values averaged across the whole electrode grid), B) MUAP duration values (half-length of the MUAP duration averaged across the whole electrode grid). Significant difference between groups, p<0.05*.

Representative results of motor unit firing activity and MUAP morphology from a child with a normal electrophysiology test, as well as from a child with electrophysiological findings suggestive of myopathy, are shown in **Figure 6**.

**Figure 6.**
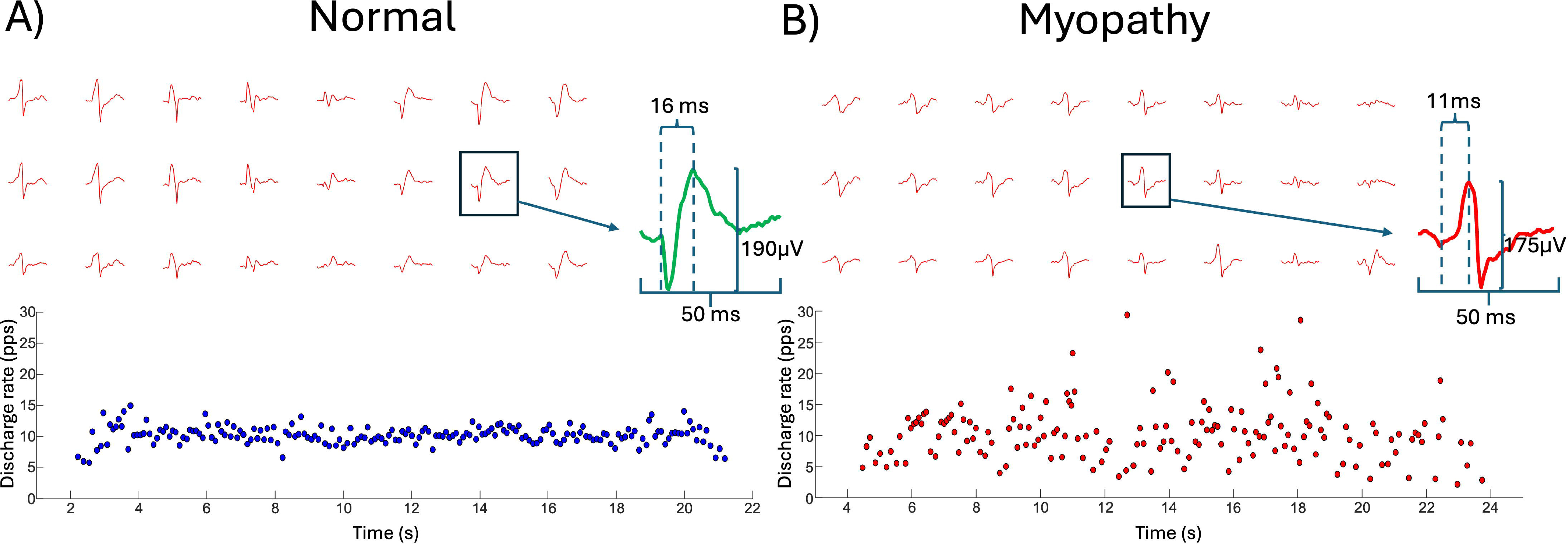
Representative MUAP spatial profiles and motor unit discharge patterns. A) individual with normal electrodiagnostic impression. MUAP profile is displayed above (24 channels). A single representative MUAP (green) with its half duration and peak to peak amplitude values can be seen on the right. Motor unit firing pattern of this motor unit can be seen below. When considering all 59 bipolar channels and the full pool of identified motor units, average MUAP half duration and peak to peak amplitude for this participant was 11.9 ms and 175.7 µV, respectively. B) individual with myopathy diagnostic impression. MUAP profile is displayed above (24 channels). A single representative MUAP (red) with its half duration and peak to peak amplitude values can be seen on the right. Motor unit firing pattern of this motor unit can be seen below. When considering all 59 bipolar channels and the full pool of identified motor units, average MUAP half duration and peak to peak amplitude for this participant was 14 ms and 190.2 µV, respectively. Despite the differences in MUAP amplitude and duration between these participants it is important to note the great variability in MUAP morphology across the electrode grid. Furthermore, note the more variable firing pattern for the individual with myopathy at the bottom right of the figure.

## Discussion

This study demonstrates that HDsEMG is both feasible and well-tolerated in children within a clinical setting. Motor unit firings were successfully decomposed in the majority of recordings. Moreover, HDsEMG decomposition enabled a comprehensive assessment of motor unit firing patterns, suggesting that children with myopathies may exhibit greater firing variability compared to those with normal or neuropathic electrophysiology findings. Finally, MUAP duration on HDsEMG significantly correlated with CNEMG recordings, supporting HDsEMG as a potential non-invasive method for MUAP analysis in children.

### Association between CNEMG and HDsEMG

One aim of this study was to compare findings from CNEMG with those from HDsEMG. Specifically, we focused on two classic electrodiagnostic parameters: MUAP amplitude and duration. Our analysis revealed no association between MUAP amplitude measurements obtained from CNEMG and HDsEMG. Several factors likely contribute to this discrepancy, including volume conduction effects, the low pass filtering effect of subcutaneous and adipose tissue, and the increased distance between recording electrodes and muscle fibers ^19^. These influences can alter MUAP size and potentially limit its diagnostic utility for neuromuscular pathologies. In contrast, CNEMG—due to its higher selectivity—captures near-fiber potentials more effectively, allowing for a more precise assessment of MUAP morphology ^20^. Despite the lack of association in amplitude, we observed a significant correlation between MUAP half-duration (defined as the interval between the first negative turn and peak) measured across the HDsEMG grid (59 bipolar channels) and MUAP duration obtained from CNEMG. Surface EMG action potentials inherently lose many characteristics of near-fiber potentials, making it more difficult to detect polyphasic potentials, jitter, or jiggle effects. However, despite the low-pass filtering effect of tissues, certain MUAP morphological features—particularly duration—appear to be preserved in HDsEMG recordings. Due to challenges in accurately determining full MUAP duration across the entire electrode grid—especially during the repolarization and refractory phases—we opted to quantify the time between the first negative turn and the MUAP peak. This approach provided a more reliable estimate of MUAP duration. Using this metric, we observed clear reductions in MUAP duration in the myopathy group, a finding consistent with conventional electrodiagnostics. This result highlights the potential of HDsEMG as a viable alternative to CNEMG in specific clinical contexts. However, we did not observe prolonged MUAP duration in the neuropathic CNEMG group, a common characteristic of this condition. This may reflect a relatively small sample size in this group. It is also important to note that while MUAP duration measured from HDsEMG was associated with CNEMG-derived values, this correlation was weak (r = 0.3). This may be due to the substantial variability in MUAP morphology across the HDsEMG grid (see Figure 6), where channels capturing smaller, less representative MUAPs could influence both amplitude and duration estimates. Future studies should aim to refine MUAP selection criteria by focusing on the most representative action potentials for a given motor unit (e.g., those with RMS amplitudes exceeding 70% of the maximum RMS amplitude across the grid ^21^). Additionally, more sophisticated analyses incorporating spectral content assessments and artificial intelligence techniques ^22^ could further enhance the ability to distinguish pathological changes in MUAP waveforms. Despite these limitations, our findings suggest MUAPs recorded via HDsEMG could have potential diagnostic utility. This diagnostic potential could be further improved through advanced signal processing techniques designed to extract key pathological features from HDsEMG recordings.

### Benefits of HDsEMG over CNEMG

CNEMG recordings have been the standard electrodiagnostic method in children since the 1980s ^23^. Due to its high selectivity and ability to isolate individual MUAPs, CNEMG provides valuable insights into MUAP morphology. Both the size and duration of action potentials can help identify anomalies in muscle fiber conduction, which are often associated with myopathies and neuropathies. However, CNEMG presents several challenges when used in paediatric patients, as previously highlighted.

In contrast, as demonstrated in the present study, HDsEMG is significantly better tolerated and allows for the recording of larger muscle regions. Previous studies using HDsEMG to assess neuromuscular function in children have primarily relied on spatial analysis techniques to interpret interference EMG signal patterns ^11^. However, these methods are limited by common challenges associated with sEMG signals, including crosstalk, volume conduction effects, and amplitude cancellation, among other influencing factors ^12^. As a result, the discriminative power of spatial analysis techniques used with HDsEMG remains constrained. Nevertheless, the high spatial sampling capability of HDsEMG, when combined with blind-source separation techniques, enables the identification of individual motor unit firing potentials ^14,24^. This facilitates a more detailed investigation of motor unit firing dynamics during contractions, opening new possibilities for analysing motor unit function. To our knowledge, this is the first study to utilize HDsEMG motor unit decomposition to assess motor unit activity in children with neuromuscular disorders.

In this study, we successfully detected motor units with complete firing profiles in 86 out of the 100 children assessed, with an average of 7 motor units per participant. Although quantitative MUAP analysis using CNEMG often reports larger samples of motor units per contraction (8 to 14 motor units in this study, see Table 1), it is important to note this technique may average the MUAP template from a minimum of 5 MUAPs to differentiate active motor units. This limited number of discharges could restrict the ability to study motor unit firing activity during contractions. Consequently, when quantitative MUAP analysis is used in electrodiagnostic studies in children, the primary focus is assessment of MUAP morphology, reflecting changes in muscle fiber membrane depolarization rather than alterations in alpha motoneuron firing properties. In contrast, HDsEMG allows for both the assessment of motor unit firing properties and changes in MUAP morphology, thereby increasing the number of parameters that can be used to detect the presence of neuromuscular disorders.

### Motor unit firing activity and the detection of myopathies

In terms of motor unit firing properties, the motor unit populations assessed across all groups were comparable, as there were no significant differences in recruitment and de-recruitment thresholds. Mean motor unit discharge rates were also similar among groups. The assessment of motor unit firing properties in neuromuscular disorders is rare, particularly in children. Among the few studies conducted in adults, findings have been inconsistent. For instance, ^25^ reported increased discharge rates in myopathies but decreased rates in central disorders such as ALS, whereas ^26^ found no significant differences between conditions—consistent with the present study. One possible explanation for these discrepancies is the challenge of accurately tracking the same motor units during contractions using earlier methodologies. If a motor unit is not consistently identified, additional motor unit activity may be erroneously merged, leading to an artificial increase in discharge rates. This issue is particularly relevant in myopathies, where motor units tend to be smaller and recruitment strategies may differ to compensate for muscle weakness ^27^. However, assessing motor unit recruitment remains challenging with any method, as motor unit identification depends largely on the volume of detectable motor unit activity within the electrode’s recording field. In the present study, the group with the lowest number of identified motor units was the myopathy group. This can likely be attributed to the characteristics of the motor unit decomposition algorithm, which may be biased to motor units with larger action potentials ^14^ while discarding those with smaller potentials—more common in myopathies. Thus, recruitment findings should be interpreted with caution, as they are influenced by both physiological factors and methodological limitations.

One of the most intriguing findings of this study was the increased variability in motor unit discharge rates observed in the myopathy group. Previous research using CNEMG has reported conflicting evidence regarding motor unit discharge variability in both neuropathic and myopathic conditions. For example, studies in adults have shown no change in firing variability in either condition ^26^ or increased variability only in neuropathic disorders ^25^. These discrepancies may stem from methodological limitations, particularly in the ability to detect and track a sufficient number of firings from a single motor unit. Many earlier studies relied on signal processing technologies that are now outdated, with most being conducted over 30 years ago. In contrast, our study allowed for the monitoring of a significant number of motor units and their firing behavior during contractions, revealing a clear increase in inter-spike interval (ISI) variability in children in our myopathy group. To our knowledge, this is the first study to report such differences in this population.

Several mechanisms may explain this finding. Potential contributors include impaired motor neuron excitability ^28^, reduced motor unit recruitment stability ^27^, and impaired neuromuscular transmission ^29^. However, given that myopathies primarily affect muscle fiber properties rather than motor neuron function, changes in motor neuron excitability are unlikely to be the primary driver of increased firing variability. Instead, altered motor unit recruitment patterns may play a key role, as individuals with myopathies often exhibit reduced motor unit innervation territories due to muscle fiber degeneration. This could lead to increased recruitment or spontaneous firing of the same motor unit in an attempt to maintain force output. Additionally, deficits in neuromuscular transmission may further contribute to ISI variability. Acetylcholine receptor dysfunction at the neuromuscular junction can lead to failed or inconsistent transmission of action potentials, disrupting motor unit firing patterns ^30^.

### Limitations

There are some limitations to this study that should be considered. First, the analysis of HDsEMG was based on the interpretation of CNEMG electrophysiological findings rather than confirmed clinical diagnoses. However, this does not impact the study’s primary aims, which were to evaluate the feasibility of using HDsEMG in a pediatric population and explore its potential as a diagnostic tool. CNEMG remains the gold standard for electrophysiological diagnosis in pediatric populations, making it a valid reference for this study. Additionally, to enhance comparability, most recordings were conducted on the tibialis anterior muscle. While this approach ensures consistency, future studies should assess multiple muscles, particularly in conditions that affect muscle groups differently (e.g., muscular dystrophies). Expanding the range of muscles analyzed would provide a more comprehensive understanding of HDsEMG’s diagnostic potential across various neuromuscular disorders. Lastly, in children capable of following instructions, surface EMG was used as feedback to regulate contraction force levels, as direct force quantification was not feasible in the clinical setting where measurements were taken. Given the high variability of surface EMG in assessing maximal contraction intensity and force control, the use of a dynamometer would have provided more precise force measurements, ensuring consistency across participants. Future studies should incorporate both HDsEMG and force measurements to better understand how motor unit function influences force generation in this population.

### Conclusion

HDsEMG motor unit (MU) decomposition is both feasible and well-tolerated in children in a clinical setting, offering valuable insights into MU firing patterns and changes in MUAP morphology. This feasibility study indicates HDsEMG recordings could be of diagnostic utility in children with neuromuscular disorders, Notably, increased MU discharge rate variability and reduced MUAP duration are identified as potential indicators of myopathic disorders, although further work is required to confirm this finding. While this study represents the first attempt to apply HDsEMG decomposition in a pediatric population, further advancements in this methodology also have the potential to enhance the diagnosis and monitoring of pediatric neuromuscular conditions.

## Author contributions

EM-V and AL designed the experiment, EM-V and AL prepared the materials, AL collected the data, EM-V, RS and IC-H processed and analyzed the data, EM-V, AL and FN interpreted the results, EM-V wrote the manuscript, EM-V, AL and FN edited the manuscript.

## Acknowledgements

This research was supported by a Muscle Dystrophy UK (MDUK) pilot data award to Eduardo Martinez-Valdes and Andrew Lawley (Award 22GRO-PG12-0572UM). Francesco Negro is funded by the European Research Council Consolidator Grant INcEPTION (contract no.: 101 045 605).

## Data availability

The data and code used in this study will be made available upon reasonable request.

